# Performance of a point of care test for detecting IgM and IgG antibodies against SARS-CoV-2 and seroprevalence in blood donors and health care workers in Panama

**DOI:** 10.1101/2020.09.25.20201459

**Authors:** Alcibiades Villarreal, Giselle Rangel, Xu Zhang, Digna Wong, Gabrielle Britton, Patricia L. Fernandez, Ambar Pérez, Diana Oviedo, Carlos Restrepo, María B. Carreirra, Dilcia Sambrano, Gilberto A. Eskildsen, Carolina De La Guardia, Yamitzel Zaldivar, Danilo Franco, Sandra López-Vergès, Dexi Zhang, Fangjing Fan, Baojun Wang, Xavier Sáez-Llorens, Rodrigo DeAntonio, Ivonne Torres-Atencio, Fernando Diaz Subía, Eduardo Ortega-Barria, Rao Kosagisharaf, Ricardo Lleonart, Chong Li, Amador Goodridge, COVID-19 serology collaborator group

## Abstract

Novel severe acute respiratory syndrome coronavirus 2 (SARS-CoV-2) is the etiologic agent of the ongoing coronavirus disease 2019 (COVID-19) pandemic, which has reached 28 million cases worldwide in eight months. The serological detection of antibodies against the virus will play a pivotal role in complementing molecular tests to improve diagnostic accuracy, contact tracing, vaccine efficacy testing and seroprevalence surveillance. Here, we aimed first to evaluate a lateral flow assay’s ability to identify specific IgM and IgG antibodies against SARS-CoV-2 and second, to report the seroprevalence of these antibodies among health care workers and healthy volunteer blood donors in Panama. We recruited study participants between April 30^th^ and July 7^th^, 2020. For the test validation and performance evaluation, we analyzed serum samples from participants with clinical symptoms and confirmed positive RT-PCR for SARS-CoV-2, and a set of pre-pandemic serum samples. We used two by two table analysis to determine the test sensitivity and specificity as well as the kappa agreement value with a 95% confidence interval. Then, we used the lateral flow assay to determine seroprevalence among serum samples from COVID-19 patients, potentially exposed health care workers, and healthy volunteer donors. Our results show this assay reached a positive percent agreement of 97.2% (95% CI 84.2-100.0%) for detecting both IgM and IgG. The assay showed a *kappa* of 0.898 (95%CI 0.811-0.985) and 0.918 (95% CI 0.839-0.997) for IgM and IgG, respectively. The evaluation of serum samples from hospitalized COVID-19 patients indicates a correlation between test sensitivity and the number of days since symptom onset; the highest positive percent agreement (87% (95% CI 67.0-96.3%)) was observed at ≥15 days post-symptom onset. We found an overall antibody seroprevalence of 11.6% (95% CI 8.5-15.8%) among both health care workers and healthy blood donors. Our findings suggest this lateral flow assay could contribute significantly to implementing seroprevalence testing in locations with active community transmission of SARS-CoV-2.

## INTRODUCTION

Coronavirus disease 2019 (COVID-19) is a viral pneumonia and multi-systemic disease caused by severe acute respiratory syndrome coronavirus 2 (SARS-CoV-2), which first appeared in Wuhan, China in December 2019 [1, 2]. Since then, the virus has spread rapidly with an explosive increase in cases across the globe. As of 12 September 2020, there have been 28,329,790 confirmed cases of COVID-19, including 911,877 deaths reported to WHO [3]. According to the Pan American Health Organization (PAHO), by August, Panama had a rate of 1,618 infected persons per 100,000 inhabitants [4, 5], placing it as the country with the second highest rate of infection in the Americas. This high rate is in part related to the fact that Panama is among the countries in the region that have conducted the highest number of tests. More than 250,000 molecular tests have been conducted since early March, and daily positivity rates have consistently been in the 30% range; there have been more than 100,000 confirmed cases. In Panama, the majority of reported cases (93.2%) demonstrate mild symptoms, while 6.8% have required hospitalization. To date, there have been 2,140 deaths and over 72,000 patients have recovered [6].

Until a vaccine becomes available, most countries’ containment efforts have relied heavily on non-pharmacological interventions to mitigate and suppress the disease. These include, but are not limited to, movement restrictions and reduced individual contact to decrease community transmission [7]. As a result, the COVID-19 pandemic, in addition to being a public health emergency, has become a financial and sociopolitical crisis. Consequently, public health strategies are urgently needed in order to ease lock-down restrictions [8]. One of the most effective strategies includes prompt and accurate diagnosis. The development of a diagnostic test that can be scaled-up to allow for mass screening among specific high-risk groups, such as health care workers (HCW), remains a key step [9, 10]. Such a test would aid with diagnosis, contact tracing, and vaccine evaluation, while also allowing the release of individuals from quarantine, as well as serological surveillance at the local, regional, and national level [11].

More than 150 diagnostics tests have been developed since the beginning of the COVID-19 pandemic [12]. The most-used platforms are enzyme-linked immunosorbent assays (ELISA) and rapid lateral flow immunoassays (LFIA) [13]. In general, serological tests based on an LFIA platform are cost- and time-efficient, do not require sophisticated equipment or highly trained personnel, and can be used to assess population exposure. In addition, LFIA platforms have generated substantial interest because they are cheaper to manufacture, store, and distribute, and easier to implement as a point-of-care test. Unfortunately, some countries have rushed into large-scale deployment of rapid tests but have found that the clinical sensitivities are low and of poor value due to inadequate performance assessment [14,15]. The performance of rapid tests provided by manufacturers might show variations. Consequently, rapid tests should be rigorously validated in a large target population before being used as a stand-alone screening test [16].

Our study aimed to evaluate the performance of an LFIA for the detection of IgM and IgG anti-SARS antibodies in COVID-19-positive individuals[17]. First, we determined the test performance of an LFIA as a rapid serology test, using a standard panel of sera from COVID-19 patients and pre-pandemic donors. Second, we conducted a field evaluation of the LFIA test to determine the seroprevalence of anti-SARS-CoV-2 antibodies among healthy blood donors (HD) and health care workers (HCW). We found this test to be suitable for conducting seroprevalence studies, assessing population exposure to the virus, and for evaluating the effects of lock-down flexibilization strategies.

## MATERIALS AND METHODS

### Lateral flow immunoassay overview

We obtained an LFIA test developed by Dr. Chong Li’s group from Institute of Biophysics, Chinese Academy of Science. The qualitative test (referred to as CAST – *Chinese Academy of Science Test*, from this point) detects and is capable of differentiating between specific IgM and IgG antibodies against SARS-CoV-2. The CAST uses a colored conjugate pad containing a recombinant SARS-CoV-2 nucleocapsid protein conjugated with colloid gold as the antigen. The CAST manufacturer’s calculated analytical sensitivity and specificity for both IgM and IgG anti-SARS-CoV-2 antibodies at development were 87.01% and 98.89%, respectively. During development in China, no cross-reactivity was reported with specimens from patients infected with Human Immunodeficiency Virus (HIV), Hepatitis A Virus (HAV), Hepatitis B soluble Antigen, Hepatitis C Virus, *Treponema pallidum*, Human T Lymphocyte Virus (HTLV), Cytomegalovirus, Influenza Virus type A and B, Respiratory Syncytial Virus, Human Papilloma Virus, *Chlamydophila pneumoniae, Legionella pneumophila, Mycoplasma pneumoniae*, or Human Parainfluenza viruses, as well as other coronaviruses. Moreover, no cross-reactivity or interference was observed with endogenous substances, including common serum components, such as lipids, hemoglobin, bilirubin, albumin, uric acid, and glucose, or other common biological analytes, such as acetaminophen, acetoacetic acid, benzoylecgonine, caffeine, EDTA, ethanol, gentisic acid, β-Hydroxybutyrate, methanol, phenothiazine, phenylpropanolamine, and salicylic acid.

### Test performance evaluation by national reference laboratory

In Panama, the CAST was evaluated independently by the Gorgas Memorial Institute of Health Studies (GMI), the National Reference Public Health laboratory responsible for COVID-19 diagnostic test validation, as well as for national molecular SARS-CoV-2 diagnoses. The real-time reverse-transcription-polymerase chain-reaction (RT-qPCR) assay for detecting SARS-CoV-2 was used as a non-reference standard [18]. We included 53 SARS-CoV-2 positive and negative samples confirmed by RT-qPCR and 55 pre-pandemic samples that included sera from patients with Dengue and Tuberculosis, in total 108 samples (Figure 2). All RT-qPCR assays were performed at GMI by trained technicians following best clinical laboratory practices and quality control assurance programs. The diagnostic accuracy of the CAST was evaluated as indicated below in the statistical analysis section.

### Field evaluation of CAST: Study participants and sample distribution

This study was conducted between April 30^th^ and July 7^th^, 2020 in four private and public hospitals and two donation centers located in Panama and Colon cities (Figure 1). The sample size was calculated using an estimated sensitivity of at least 80% and a specificity of at least 90% for the CAST. Based on the target population of the study, which included positive cases and contacts, we assumed a prevalence of at least 15%. Thus, the sample size was estimated at a minimum of 650 participants, aiming for a 95% level of accuracy. The inclusion criteria were being an adult over 18 years old and providing written informed consent. All study participants completed a clinical screening survey for COVID-19-related symptoms and consented to submit samples for screening of other infections. Only healthy blood donors (HD) that tested negative for other infectious diseases, including Chagas disease, HIV, HBV, HAV, and HTLV1, were invited to participate in our study. The exclusion criteria comprised those with missing data and patients in intensive and semi-intensive care units. Health care workers (HCW) were asked to provide an additional blood sample 15 days after the first sample was taken. All HCW in contact with confirmed COVID-19 cases were considered high risk. This study was registered with the Panama Ministry of Health (No. 1462) and was approved by the National Research Bioethics Committee (CNBI; No. EC-CNBI-2020-03-43).

**Figure 1.**
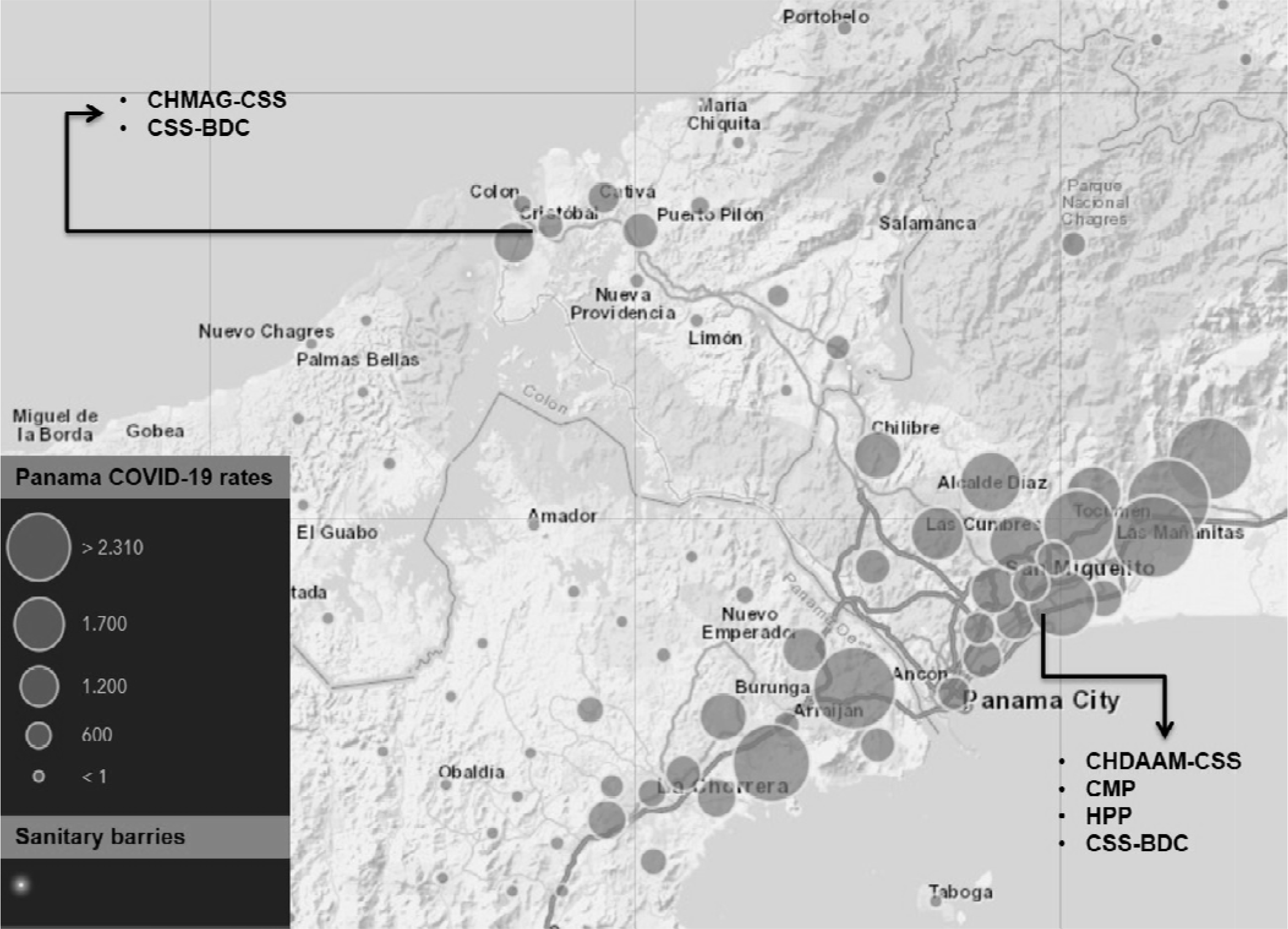
Panama study sites and health institutions involved. Gray circles on the geographical map of Panama indicate the number of active COVID-19 cases in the areas with the highest incidence of reported cases. Input data June 2020. Source: Panama Ministry of Health. Black arrows indicate the locations of patients and health care workers recruited for this study. The sites included two private hospitals and two public hospitals in Panama and Colón cities. CMP=Centro Médico Paitilla; HPP=Hospital Pacífica Salud; CHDAAM-CSS= Complejo Hospitalario Dr. Arnulfo Arias Madrid-Caja de Seguro Social; CHAMAG-CSS= Complejo Hospitalario Manuel Amador Guerrero-Caja de Seguro Social; CSS-BDC= Caja de Seguro Social Blood Donor Centers.

### Specimen collection, demographic and clinical data

Predesigned questionnaires related to COVID-19 from the World Health Organization (WHO) were completed by trained interviewers. This questionnaire was given to all study participants. Information on sociodemographic factors, medical history, current COVID-19 symptoms, and epidemiological data were collected through personal interviews. Questions related to anosmia and ageusia symptoms were not included in the interview. Venous blood samples were collected from all the study participants for CAST analysis. The blood collection tubes were kept at room temperature to allow clot formation and then centrifuged for 10 min at 250 x g to obtain serum specimens. All rapid test analyses were conducted with fresh serum samples.

### IgM and IgG antibody detection by LFIA

We followed a step-wise protocol for conducting the CAST. Briefly, we added one drop of serum (approximately 20-25 μl) into the cassette sample well followed by two drops of the provided buffer (approximately 70 μl). If IgM and/or IgG anti-SARS-CoV-2 antibodies are present in the sample, they will bind to the colloidal gold conjugate, forming an immunocomplex. This immunocomplex is then captured by the respective pre-coated band containing either anti-IgM or anti-IgG antibodies, forming a red colored IgM and/or IgG line. The presence of one red lines indicates the sample is positive for specific IgM or IgG anti SARS-VoV-2 antibodies, while the presence of two lines indicates the sample is positive for both IgM and IgG antibodies. A third line functions as a positive control, indicating that the kit is working properly. All analyses were interpreted by two independent technicians at 15 minutes after the serum was added. If there were disagreements, a third trained technician evaluated the result and provided the final decision.

### Statistical analysis

Data were analyzed with SPSS version 25.0 (Armonk, NY: IBM Corp.). A descriptive analysis was performed to calculate the frequencies and percentages for categorical variables. Continuous variables were presented as the mean ± standard deviation (SD). For the groups evaluated with and without COVID-19 disease, the rapid test results were compared against the non-reference standard RT-qPCR. Estimations of *Kappa* and positive percentage agreement (PPA) were calculated with a 95% confidence interval [19]. *P*-values <0.05 were considered statistically significant.

## RESULTS

### Study site and study participants

This study was conducted between April 30^th^ and July 7^th^, 2020 in four private and public hospitals located in Panama and Colon cities, as well as the blood donation center in Panama City (Figure 1). A total of 702 participants were recruited for the field study: 255 (36.3%) were HD, while the remaining 63.7% of the sample comprised 351 HCW and 96 COVID-19 patients (confirmed by RT-qPCR) (Figure 2). **Table 1** summarizes the age, sex, COVID-19 exposure, and presence of comorbidities across participants in the COVID-19 patient, HCW, and HD groups. Among the participants from COVID-19 group, 67 (69.9%) reported a pre-existing chronic disease; whereas 90 (25.6%) HCW and 28 (11.0%) HD reported a pre-existing chronic disease.

**Figure 2.**
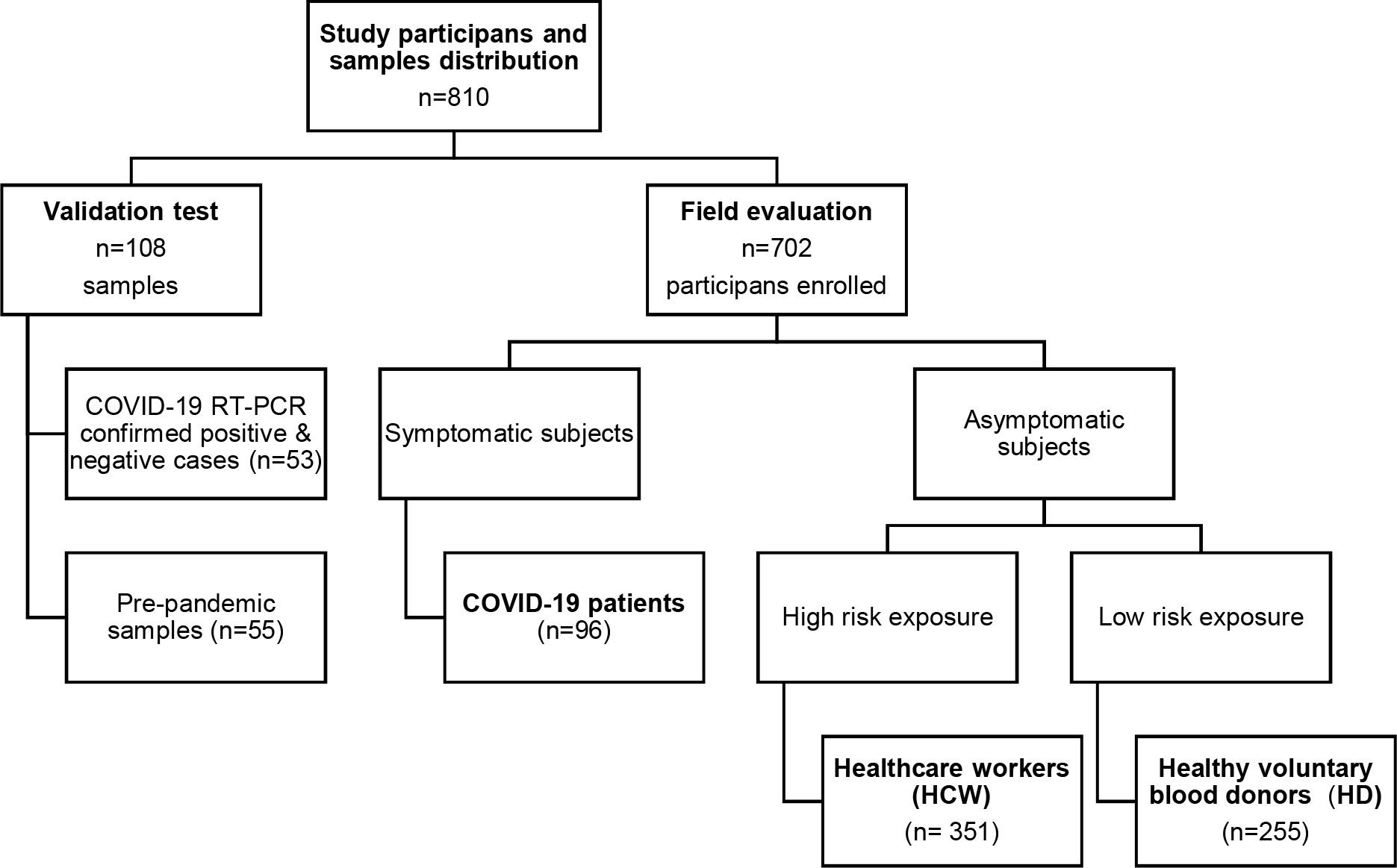
Schematic representation of study participants and samples distribution. A total of 810 blood samples for detection of IgM and IgG anti SARS-CoV2 antibodies were analyzed for this study. Performance tests included COVID-19 RT-PCR confirmed +/- cases (n=53) and a pre-pandemic panel of samples (n=55). For the field study evaluation a total of 702 participants were enrolled, classified as follows: COVID-19 patient (n=96), Health care workers (n= 351) and Healthy volunteer blood donors (n=255).

**Table 1.** Sociodemographic and comorbid information of study groups according the positive CAST results.

|  | COVID-19 patients<br>Mean (SD) or # (%) |  | Healthcare workers (HCW)<br>Mean (SD) or # (%) |  | Healthy voluntary donors (HD)<br>Median (SD) or # (%) |  |
| --- | --- | --- | --- | --- | --- | --- |
| Variables | Total participants<br>(n=96) | CAST Positive<br>(n= 65) | Total participants<br>(n=351) | CAST Positive<br>(n=45) | Total participants<br>(n=255) | CAST Positive<br>(n= 34) |
| **Age** | 54.9 (15.7) | 53.1 (14.5) | 39.5 (11.5) | 41.1 (11.3) | 73.2 (11.1) | 35.5 (10.3) |
| **<20** | 0 (0%) | 0 (0%) | 0 (0%) | 0 (0%) | 7 (2.7%) | 1 (2.9%) |
| **20-39** | 16 (16.7%) | 10 (15.4%) | 198 (56.4%) | 22 (48.9%) | 141 (55.3%) | 22 (64.7%) |
| **40-59** | 44 (45.8%) | 35 (53.8%) | 137 (39.0%) | 19 (42.2%) | 100 (39.2%) | 11 (32.3%) |
| **60-79** | 31 (32.3%) | 17 (26.1%) | 19 (5.4%) | 4 (8.9%) | 7 (2.7%) | 0 (0%) |
| **>80** | 5 (5.2%) | 3 (4.6%) | 0 (0%) | 0 (0%) | 0 (0%) | 0 (0%) |
| **Sex** |  |  |  |  |  |  |
| **Male** | 64 (66.7%) | 46 (70.8%) | 113 (32.3%) | 10 (22.2%) | 181 (71.0%) | 23 (67.6%) |
| **Female** | 32 (33.3%) | 19 (29.2%) | 238 (67.8%) | 35 (77.7%) | 74 (29.0 %) | 11 (32.4%) |
| **COVID-19 contact** |  |  |  |  |  |  |
| **No contact** | 27 (29.0%) | 15 (23.1%) | 52 (14.8%) | 5 (11.1%) | 222 (87.0%) | 24 (70.6%) |
| **Contact** | 34 (36.6%) | 26 (40.0%) | 256 (72.9%) | 34 (75.6%) | 20 (7.8%) | 7 (20.6%) |
| **Doesn't know** | 32 (34.4%) | 23 (35.4%) | 43 (12.2%) | 6 (13.3%) | 13 (5.1%) | 3 (8.8%) |
| **Chronic diseases** |  |  |  |  |  |  |
| **Hypertension** | 46 (47.9%) | 33 (50.8%) | 55 (15.7%) | 6 (13.3%) | 15 (5.9%) | 1 (2.9%) |
| **Renal failure** | 13 (13.5%) | 6 (9.2%) | 1 (0.3%) | 1 (2.2%) | 0 (0%) | 0 (0%) |
| **Respiratory insufficiency** | 3 (3.1%) | 1 (1.5%) | 0 (0%) | 0 (0%) | 0 (0%) | 0 (0%) |
| **Cardiac insufficiency** | 3 (3.1%) | 2 (3.1%) | 0 (0%) | 0 (0%) | 0 (0%) | 0 (0%) |
| **Cancer** | 5 (5.2%) | 2 (3.1%) | 0 (0%) | 0 (0%) | 0 (0%) | 0 (0%) |
| **Diabetes** | 29 (30.2%) | 21 (32.3%) | 18 (5.1%) | 2 (4.4%) | 0 (0%) | 0 (0%) |
| **Asthma** | 1 (1.0%) | 1 (1.5%) | 11 (3.1%) | 4 (8.9%) | 7 (0%) | 1 (2.9%) |
| **Other<sup>1</sup>** | 7 (7.3%) | 6 (9.2%) | 23 (6.5%) | 3 (6.7%) | 5 (0%) | 2 (5.8%) |

**Notes:** <sup>1</sup>other chronic diseases reported by participants were dyslipidemia and thyroid disease. COVID-19 contact refers to those participants that had contact with a confirmed COVID-19 case in the household and/or during daily activities at work in a healthcare facility.

### CAST test diagnostic performance using panel of reference sera

Samples including positive and negative COVID-19 cases confirmed by RT-PCR, and a set of pre-pandemic panel samples were analyzed with the CAST device. A comparison of the CAST and RT-PCR results and the analytical performance results are shown in Table 2. For both IgM and IgG antibodies, the test demonstrated a negative predictive value (NPV) of 0.985% (95% C.I. 0.915-1). An evaluation of the negative percent agreement indicated that of the cases with negative RT-PCR test results, only five showed positive CAST results for IgM and four showed positive CAST results for IgG. Two of these cases that were positive for both IgM and IgG were later determined to be false RT-PCR negatives based on two additional commercial lateral flow immunoassays (data not shown). Virus clearance may explain the other cases that showed negative RT-PCR results but positive antibodies results with the CAST. Of the pre-pandemic samples tested (n=55), only one from a patient with a positive result for Dengue showed a positive IgM result on the CAST platform (data not shown).

**Table 2.** Diagnostic performance and diagnostic certainty of CAST using a panel of reference sera.

| CAST device |  | RT- PCR SARS-CoV-2 |  |  | Positive<br>percent<br>agreement<br>(PPA)<br>(95% CI) | Negative<br>percent<br>agreement<br>(NPA)<br>(95% CI) | Overall<br>percent<br>agreement<br>(95% CI) | Kappa<br>(95% CI) |
| --- | --- | --- | --- | --- | --- | --- | --- | --- |
|  |  | Positive | Negative | Total |  |  |  |  |
| CAST IgM | Positive | 35 | 4 | 39 | 97.2%<br>(84.6-100.0) | 94.4%<br>(86.2-98.2) | 95.4%<br>(89.3-98.3) | 0.898<br>(0.811-0.985) |
|  | Negative | 1 | 68 | 69 |  |  |  |  |
|  | Total | 36 | 72 | 108 |  |  |  |  |
| CAST IgG | Positive | 34 | 4 | 38 | 97.2%<br>(84.6-100.0) | 95.8%<br>(88.0-99.1) | 96.3%<br>(90.6-98.9) | 0.918<br>(0.839-0.997) |
|  | Negative | 0 | 70 | 70 |  |  |  |  |
|  | Total | 34 | 74 | 108 |  |  |  |  |

We proceeded to evaluate the CAST’s performance in the field during the current COVID-19 pandemic in Panama. We recruited 96 COVID-19 ward patients (Figure 2). All participants from this group were RT-PCR-confirmed positive cases and developed moderate COVID-19 symptoms. Analysis of the COVID-19-confirmed patient group showed a PPA of 67.7% (95% CI 57.8-76.2%) for IgM and IgG anti-SARS-CoV-2 antibodies (Data not shown). In order to investigate seroconversion over the course of COVID-19 evolution in patients, the data from 66 sera samples were divided into three groups according to the time of sample collection after illness onset. The CAST results showed a PPA of 36.4% (95% CI 19.6-57.1%) for either or both IgM and IgG in patients whose samples were collected from 0-7 days after RT-PCR diagnosis (Table 3). PPA scores of 76.2% (95% CI 54.5-89.8%) and 71.4% (95% CI 49.8-86.4%) for IgM and IgG, respectively, were found for patients whose samples were collected from 8-14 days after positive RT-PCR results. The highest PPA score of 87.0% (95% CI 67.0-96.3%) for both IgM and IgG antibodies was found for samples collected more than 15 days after diagnosis (Table 3).

**Table 3.** CAST SARS-CoV-2 IgM and IgG PPA by days post symptom onset in COVID-19 patients.

| Time from symptom onset, days<sup>1</sup> | Positive /total samples tested | PPA (%) | 95% CI |
| --- | --- | --- | --- |
| 0-7 | 8/22 | 36.4 | 19.6-57.1 |
| 8-14 | 15/21 | 76.2 | 54.5- 89.8 |
| ≥15 | 20/23 | 87.0 | 67.0-96.3 |

Note: <sup>1</sup>Considering RT-PCR confirmed SARS-CoV-2 cases. PPA= Positive Percent Agreement

### Field evaluation of CAST among health care workers and healthy blood donors

To determine seroprevalence among a potentially exposed population and a population of healthy donors, we applied the CAST to participants with a high (HCW) and low (HD) risk of exposure to the virus. We found that forty-five out of 351 HCW tested positive for both IgM and IgG SARS CoV-2 antibodies, which corresponds to a prevalence of 11.61% (95% CI 8.6-15.4%) (Figure 3). In contrast, 86.97% (95% CI 83.0-90.1%) of the HCW samples were non-reactive, while 0.28% (95 C.I 0-1.7%) and 1.42% (95% CI 0.5-3.4%) of the HCW samples were positive for only IgM or only IgG, respectively (data not shown). Next, we determined the seroprevalence among a group of HD and found that 11.72% (95% CI 8.3-16.3%) of the samples from this group were positive for both IgM and IgG anti-SARS-CoV-2 antibodies (Figure 3).Thus, 85.94% (95% CI 81.1-89.7%) of the HD samples were non-reactive, while 0.78% (95% CI 0-3.0%) of the HD samples were positive for only IgM or only IgG antibodies. We also found that there were no seroprevalence differences among HCW with clinical responsibilities (nurses and physicians) compared to those without clinical responsibilities (administrators, laboratory technicians, etc.) (12.9% vs. 14.1%, respectively). In order to determine the risk of exposure during interactions with hospitalized COVID-19 patients, we asked HCW if they had been in contact with those patients. We found that those HCW that reported having close contact with confirmed COVID-19 cases demonstrated a higher seroprevalence than HCW that did not report close contact (12.4% vs. 1.8%, respectively, data not shown).

**Figure 3.**
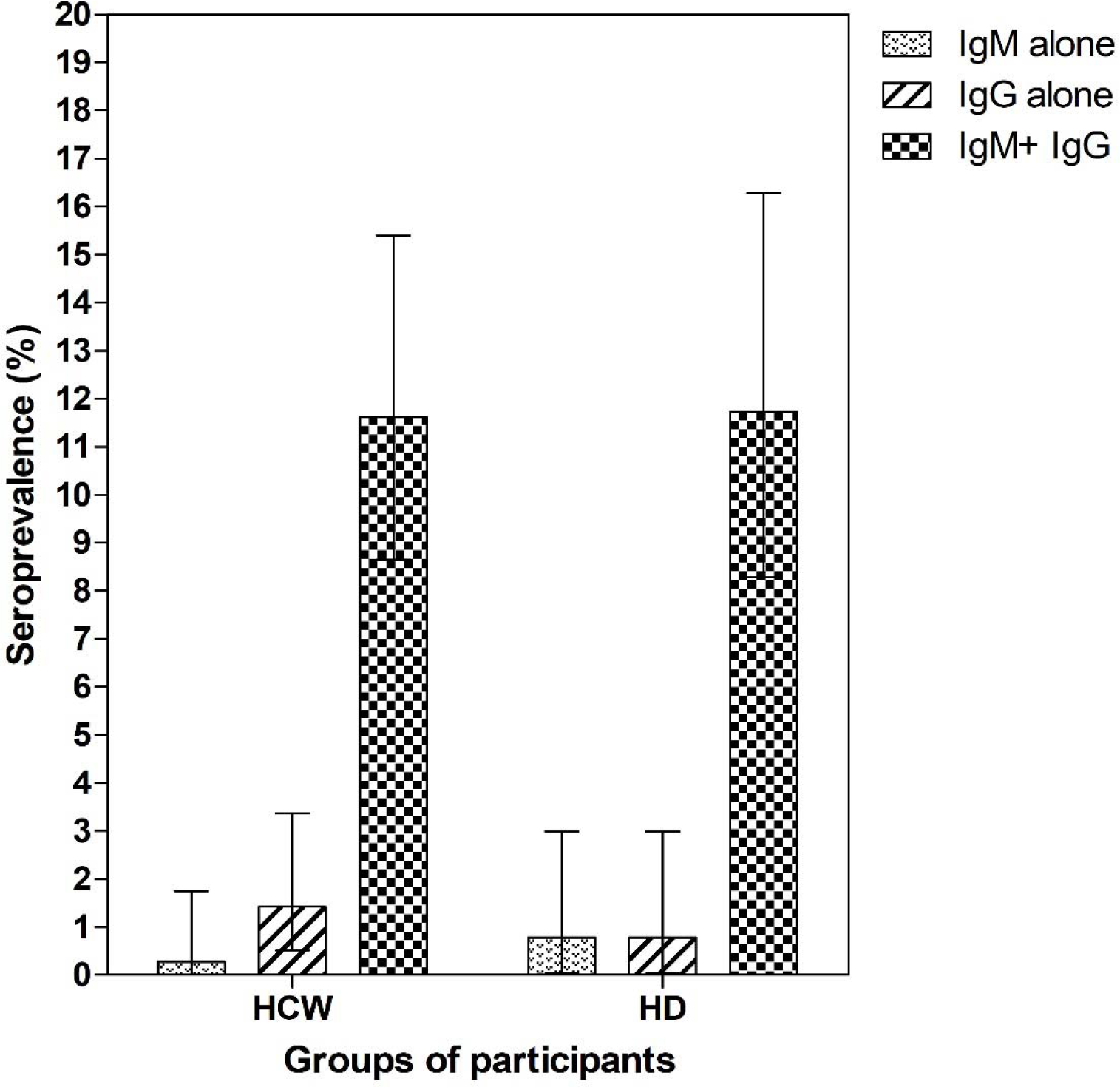
Seroprevalence of SARS-CoV2 IgM and IgG in health care workers and healthy volunteer blood donors. A total of 351 healthcare workers (HCW) and 255 healthy volunteer donors (HD) were analyzed by rapid test for detection of anti SARS-CoV2 IgM and IgG antibodies. Each bar represents seroprevalence (%) according to the detection of IgM, IgG, or both IgM and IgG antibodies. Error bars represent the 95% CI.

We also analyzed differences in the age and gender of seropositive participants. In both the HCW and HD groups, we found the highest seropositivity among participants in the age range of 20-39 years (48.9% and 64.7%, respectively). Among seropositive HCW, 77.8% were female and 22.2% male (Table 1). The majority of HCW (75.6%) reported having contact with a confirmed COVID-19 case, while most of the HD participants reported no contact (70.6%).

## DISCUSSION & CONCLUSION

Here we report COVID-19 antibody seroprevalence in HCW and HD in Panama. We also report the performance of a rapid test kit for detecting anti-SARS-CoV-2 IgM/IgG antibodies with an LFIA. We tested serum samples from confirmed positive and negative COVID-19 patients and pre-pandemic samples collected in 2019. Our analysis showed a high *Kappa* correlation, indicating very close agreement between the RT-PCR and CAST. Rashid and colleagues described several rapid detection test kits and reported sensitivities for both IgM and IgG ranging between 72.7% and 100%, while specificities ranged from 98.7% to 100% [20]. Adam *et al* estimates that the sensitivity of rapid tests for the detection of anti-SARS-CoV-2 antibodies ranged from 55-70% when compared to RT-PCR and 65-85% when compared to ELISA, with specificities of 95-100% and 93-100%, respectively [21]. Based on our results, we conclude that the CAST is suitable for seroprevalence studies.

Our study reports the seroprevalence of anti-SARS-CoV-2 IgM/IgG antibodies among a representative sample of HCW and HD in Panama. Specifically, we investigated the seroprevalence in a population of hospitalized COVID-19 patients and a group of participants with a high risk of infection (health care workers) and another group with a low risk of contagion (healthy donors).

When we stratified the COVID-19 patient samples according to when they were collected in terms of number of days after symptom onset, we observed differences in the prevalence of positive results. A positivity rate of 87.0% (95% CI 67.0-96.3%) for both IgM and IgG antibodies was found in samples collected 15 days or more after a positive RT-PCR result. Similar to our findings, Pan *et al* reported rapid test positivity rates of 11.1%, 92.9%, and 96.8% at the early convalescent (1–7 days after onset), intermediate (8–14 days after onset), and late convalescent stages (more than 15 days) of infection, respectively [22]. High sensitivity of serological testing two weeks after symptom onset has been shown in other studies [23]. In a study by Severance *et al*, 100% sensitivity was seen at ≥15 days post-PCR diagnosis, and Tang *et al* reported 93.8% sensitivity (95% CI; 82.80-98.69) at ≥14 days post-symptom onset [24, 25]. Based on our data, the usefulness of the CAST increased significantly two weeks post symptom onset. This observation indicates that the CAST could be used to evaluate the anti-SARS-CoV-2 antibody response 14 days after symptom onset, when the positivity rates are highest.

Regarding the results in HCW and HD participants, we found a seroprevalence of IgM and IgG antibodies of 11.61% (95% CI 8.6-15.4%) and 11.72% (95% CI 8.3-16.3%) in the HCW and HD groups, respectively. Given that HCW are a high-risk population [26, 27], it was quite surprising to find that the seroprevalence was nearly equal between the two groups, similar to a previous report [28]. It is worth mentioning that special precautions for blood donations are under investigation. To date, previous reports suggest no direct threat to blood safety itself [29-31]. Likewise, it is important to highlight the ability of this rapid test to detect antibodies in mild or asymptomatic COVID-19 populations since there are indications that less severe illness is associated with lower antibody titers [32-35]. As none of the members of this group reported being hospitalized or having symptoms clearly indicating recent infection, it is tempting to conclude that these represent asymptomatic cases that were infected while exposed to COVID-19 patients or in the community.

Theoretically, positive IgM and IgG tests for SARS-CoV-2 antibodies detected in patient blood samples indicate that it is likely that the individual is in the early convalescent stage of infection. Indeed, serology testing provides an important complement to RNA testing in the later stages of COVID-19 [36]. If only IgG antibodies are detected, then it is probable that the person had an infection sometime in the past or the patient is in the convalescent stage of infection. Our study reveals the positivity rates for IgM and IgG between 0 and 15 days after symptoms onset. Several scientists have reported that the detectable serology markers IgG and IgM have similar seroconversion in COVID-19 patients, with antibody levels increasing rapidly starting from 4 to 6 days after the appearance of symptoms [21, 22, 37, 38]. Comprehensive studies looking at anti-SARS-CoV-2 antibody dynamics are warranted to fully describe their dynamics in the short and long term after infection.

Our study has several limitations. First, we were not able to use samples from individuals with other respiratory tract infections to rule out cross reactivity with human coronaviruses causing common seasonal colds. However, a set of 55 pre-pandemic samples was used to validate the test, and all but one tested negative for anti-SARS-CoV-2 antibodies. Second, this rapid test is based on a colorimetric evaluation of the IgG and IgM bands determined by an operator, which implies the limitations that a qualitative inter-intra-operator evaluation might produce in terms of variability. In our study, this limitation was addressed by resorting to double operator evaluation and taking photographs of all test results to be re-analyzed by a third party in the case of first level evaluation disagreement. Third, the CAST is a qualitative detection method; thus, the antibody levels in COVID-19 patients were not measured in this study. Also, the sample size was calculated for the minimum sample required to validate the test. It is yet to be determined if the CAST produces the same results in a point-of-care setting using fresh blood samples since we used serum after centrifugation. Moreover, we have not evaluated if the CAST produces comparative results with ELISA or immunochemiluminescent tests. Ongoing work by our research team will allow us to establish the CAST’s limits of detection by comparing results with an ELISA test.

The CAST (rapid test) has some advantages compared to other more complex laboratory-based tests. Compared to automated ELISA and immunochemiluminescent assays, CAST is economical and time efficient, does not require advanced equipment, is simple to perform, and requires minimal training. The CAST can be used for seroprevalence studies in primary health care settings as well as in specific contexts outside of hospitals, such as high-prevalence areas. Due to its low cost and short turnaround time, CAST is suitable for large-scale sample screening. In addition, using blood samples as opposed to nasal swabs could eliminate the need for operational steps that may produce aerosols and place technicians at higher risk. Some groups have attempted to compare serological tests with RT-PCR platforms. These tests have different targets and applications. The RT-PCR is intended for acute phase diagnosis, while the serology tests are intended for antibody seroprevalence studies.

In conclusion, the findings of this cross-sectional study demonstrate the value of the CAST for the detection of specific IgM and IgG antibodies at the population level, including health care personnel, healthy blood donors and other community members. The use of a rapid test among both healthy individuals and patients to conduct surveillance in outbreak areas could provide critical information about the status of the COVID-19 pandemic. Such a rapid test would allow the characterization of the pandemic’s behavior at the community level and the identification of transmission hot spots in the community, which, in turn, would help us to better understand the situation and establish optimal strategies within quickly changing epidemic scenarios [15]. In addition, it will facilitate the massification of diagnostic methods allowing us to determine the seroprevalence of the Panamanian population and the true extent of SARS-CoV-2 community spread.

## Data Availability

All data will be available upon request by email to

## ACKNOWLEDGEMENTS

We thank all the patients affected by COVID-19 for their participation in our study. We also thank the health care workers from all participating hospitals for their contributions to sampling and providing key information on the case distribution network. Special thanks to Dr. Victor Sanchez from SENACYT for his support of the collaboration network with the Ministry of Health and the National COVID-19 response. Lastly, we thank Colleen Goodridge for her critical review of the manuscript and valuable suggestions. This manuscript has been released as a pre-print at MedRxiv, (Villarreal *et al*.).

---

**Supplementary table.** Symptoms among COVID-19 patients.

| Symptoms | COVID19 participant<br># (%) |
| --- | --- |
|  | **Total**<br>**(N=96)** |
| **Fever** | 64 (66.7%) |
| **Chills** | 45 (58.4%) |
| **Fatigue** | 50 (49.0%) |
| **Muscular pain** | 39 (40.6%) |
| **Sore throat** | 37 (38.5%) |
| **Cough** | 73 (76.0%) |
| **Rhinorrhea** | 29 (30.2%) |
| **Dyspnea** | 68 (70.8%) |
| **Wheezing** | 21 (21.9%) |
| **Chest pain** | 39 (40.6%) |
| **Other breathing symptoms** | 8 (8.4%) |
| **Headache** | 52 (54.2%) |
| **Nausea or vomit** | 34 (35.4%) |
| **Abdominal pain** | 29 (30.2%) |
| **Diarrhea** | 56 (58.9%) |

